# Artificial Intelligence and Mobile Health Technologies for Improved Healthcare Access: Design and Pilot Evaluation of an Integrated Digital Health Platform for a Nigerian Teaching Hospital

**DOI:** 10.64898/2026.09.02.26361014

**Authors:** Ogorchukwu Emmanuel Ochem, Okwuchukwu Praise Ezego

## Abstract

Healthcare access barriers among Nigerian university students remain under-researched, particularly regarding localized digital health interventions. This study designed, developed, and evaluated a mobile health platform integrating AI-assisted triage, teleconsultation, appointment scheduling, and records management for students at the University of Abuja Teaching Hospital (UATH). A single-group pre-post design was conducted with 50 enrolled student-patients, with each participant observed for 30 days. The platform’s triage feature utilized MedGemma, deployed locally via Ollama on UATH infrastructure using a multi-turn conversational architecture. Evaluation metrics included a survey operationalizing the Penchansky and Thomas access framework, the System Usability Scale (SUS), and triage validation against two independent clinicians using weighted Cohen’s kappa. Among 61 in-scope triage sessions from 28 participants, mean triage response time was 34.8 seconds with no performance degradation. Among 22 participants completing the SUS, the mean score was 73.2, significantly exceeding the standard 68 benchmark (p = .046). Healthcare access improved directionally across four of five measured dimensions. AI triage classifications demonstrated substantial agreement with independent clinicians **(κ = 0.66–0.72)**, strengthening in longer conversations. No significant access equity differences were detected between on-campus and off-campus participants in the underpowered pilot subgroup analysis. This study demonstrates the technical feasibility, usability, and clinical concordance of a locally deployed, AI-assisted health platform in a resource-constrained Nigerian setting, providing, to our knowledge, the first evaluation of MedGemma for AI-assisted triage among Nigerian university students and justifying a larger, fully powered follow-up study.

## Introduction

Healthcare access barriers among university students in low- and middle-income countries are well documented globally but rarely examined at the institutional level where they are actually experienced. Universal health coverage remains a distant target under Sustainable Development Goal 3, with closing the financing gap alone estimated to require $274–371 billion annually across low- and middle-income countries (Stenberg et al., 2017), and quality-of-care failures, not just access failures, account for more than half of the 8.6 million annual deaths linked to treatable conditions in these settings (Kruk et al., 2018).

Nigeria’s own access barriers are well characterised nationally, uneven workforce distribution, low insurance coverage (under 5% nationally), and system-wide mistrust rooted in equipment shortages and inconsistent care quality (Alawode & Adewole, 2021; Ogueji et al., 2023). What remains absent from the literature is any study examining these barriers at the university level specifically, despite tertiary institutions combining large residential populations with limited on-site medical capacity. A recent national systematic review of telemedicine barriers and facilitators in Nigeria, the first of its kind, identified education and training, not technology alone, as the most consistently reported facilitator of adoption (Cole et al., 2025), a finding this study’s design directly incorporates.

Artificial intelligence-assisted triage offers a plausible route to narrowing this gap, but the evidence base for its safety is mixed. Symptom-checker tools show diagnostic accuracy ranging from just 19–38% and triage accuracy that has not meaningfully improved over a five-year follow-up period internationally (Schmieding et al., 2022; Wallace et al., 2022). MedGemma, the model used in this study, carries a stronger published evidence base than typical open-source alternatives, including validation against AfriMed-QA, a Pan-African medical question-answering benchmark, but this evidence is self-reported by the model’s developer and has not been independently evaluated against Nigerian patients in a real deployment (Google, 2026; Olatunji et al., 2024). This study addresses that specific gap.

This study had five objectives: (1) design and develop an AI-assisted mobile health platform for UATH students; (2) evaluate pre-post changes in healthcare access across the five (Penchansky & Thomas, 1981) access dimensions; (3) assess platform usability; (4) validate AI triage output against independent clinical judgement; and (5) examine differences in access outcomes between on-campus and off-campus students.

## Methods

### Design

A single-group pre-post design was used, deliberately positioned as feasibility-establishing rather than a definitive efficacy trial, consistent with Murray et al.’s (2016) criteria for when a digital health intervention is and is not ready for randomised evaluation. Fifty student-patients affiliated with UATH were enrolled: 20 on-campus and 30 off-campus (departing from an original 25/25 recruitment target), 25 female and 25 male, aged 18–30, with no known physical disability.

### Platform

The platform combines a Flutter mobile client, a Python/FastAPI backend, and a MySQL database, with MedGemma 4B deployed locally via Ollama on a server physically located on UATH’s premises, directly addressing data sovereignty concerns associated with cloud-based commercial AI APIs. Patient records are cached locally for offline review; AI-assisted triage requires connectivity to reach the institutional server. Triage conversations are multi-turn, incorporate each patient’s full recorded history as context, and conclude with a three-tier urgency classification (routine, urgent, emergency) that determines routing; an emergency classification directs the patient to in-person care rather than a virtual consultation. Video consultation (ZegoCloud) is the only paid feature; triage, scheduling, and records access are free.

### Procedure

Following informed consent and a structured onboarding session (covering AI triage limitations and escalation pathways), participants used the platform for 30 days, engaging with any combination of features as their own healthcare needs arose. For triage analyses, each participant’s first logged triage session was used as the operational start of their 30-day observation period, and only sessions occurring within that participant-specific window were included in the primary system-performance and concordance analyses. Interactions occurring after this period were treated as post-study use and excluded. Baseline and post-intervention administrations of the access survey and a one-time SUS administration (restricted to participants who used AI triage) captured outcome data.

### AI validation

Every in-scope triage session was independently reviewed by two clinicians (blinded to MedGemma’s classification and to each other’s response), who assigned their own urgency classification. Weighted Cohen’s kappa was calculated for three pairings, MedGemma-Clinician A, MedGemma-Clinician B, and Clinician A-Clinician B, in aggregate and stratified by conversation length (≤3 vs ≥4 patient turns), following (Sim & Wright, 2005) guidance on kappa interpretation and sample size limitations.

### Analysis

Paired access outcomes were analysed using paired t-tests (with Wilcoxon as a non-normality fallback) with Holm-Bonferroni correction across the five dimensions and composite score. SUS was compared against the published 68 benchmark via one-sample t-test. Subgroup comparisons used independent-samples tests on change scores. Analysis was conducted in Python (SciPy, pingouin).

## Results

Of 50 enrolled participants, 49 completed baseline and 42 (84%) completed post-intervention data collection. Thirty-three participants (66%) used AI-assisted triage at least once. The system-performance and concordance analyses comprised 61 in-scope triage sessions from 28 participants during their respective 30-day observation periods; additional sessions occurring outside these periods were excluded from these analyses.

### System performance

Sixty-one in-scope triage sessions across 28 participants showed a mean response time of 34.8 seconds (SD 13.9), with no evidence of degradation across the intervention period. Response time correlated with conversation length (r=0.726, p<.001) but not with chronological position in the study.

### Access outcomes

Across the five Penchansky and Thomas dimensions and composite score, four of five dimensions improved directionally; accessibility (p=.009) and the composite score (p=.033) reached significance before correction, but neither survived Holm-Bonferroni correction (Table 1).

**Table 1.** Pre-Post Healthcare Access Outcomes.

| Dimension | Baseline M (SD) | Post M (SD) | p (raw) | p (Holm) | Cohen's d |
| --- | --- | --- | --- | --- | --- |
| Availability | 2.98 (0.53) | 3.06 (0.63) | .076 | .304 | 0.13 |
| Accessibility | 2.62 (0.58) | 2.75 (0.62) | .009 | .052 | 0.21 |
| Accommodation | 3.12 (0.59) | 3.10 (0.64) | .574 | 1.000 | -0.04 |
| Affordability | 2.45 (0.65) | 2.47 (0.71) | .703 | 1.000 | 0.03 |
| Acceptability | 3.01 (0.55) | 3.07 (0.62) | .206 | .617 | 0.09 |
| Composite | 2.84 (0.46) | 2.89 (0.46) | .033 | .164 | 0.11 |

### Usability

Among 22 participants who completed the SUS, mean score was 73.2 (SD 11.5), significantly above the 68 benchmark (t(21)=2.12, p=.046), a result robust across parametric and non-parametric sensitivity checks despite one low outlier.

### AI triage concordance

The 61 in-scope triage sessions from 28 participants were independently reviewed by both clinicians and formed the basis of the concordance analysis. Agreement was assessed using raw percentage agreement and weighted Cohen’s kappa with linear weights. Raw agreement was calculated as the proportion of sessions for which both raters assigned the same three-tier urgency classification.

MedGemma agreed with Clinician A on 55 of 61 sessions (90.2%) and with Clinician B on 57 of 61 sessions (93.4%). The two clinicians agreed with each other on 55 of 61 sessions (90.2%). Weighted kappa similarly showed substantial aggregate agreement across all three pairings: MedGemma–Clinician A κ = 0.659 (95% CI [0.33, 0.89]), MedGemma–Clinician B κ = 0.719 (95% CI [0.42, 0.93]), and Clinician A–Clinician B κ = 0.675 (95% CI [0.42, 0.88]) (Table 2).

**Table 2.**
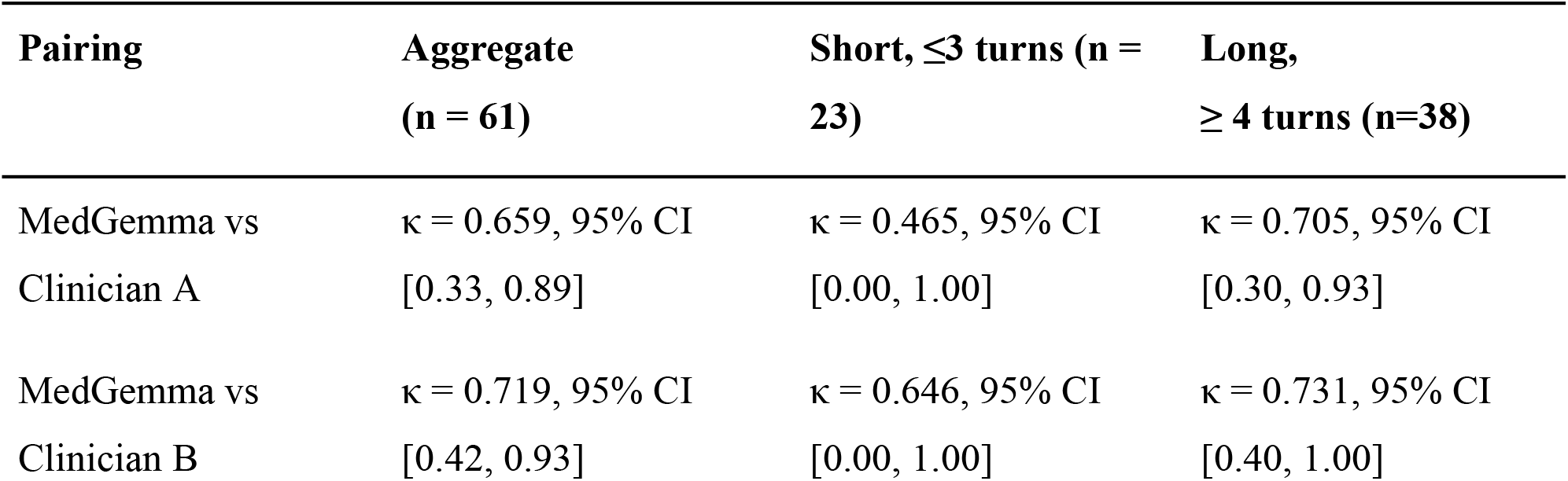
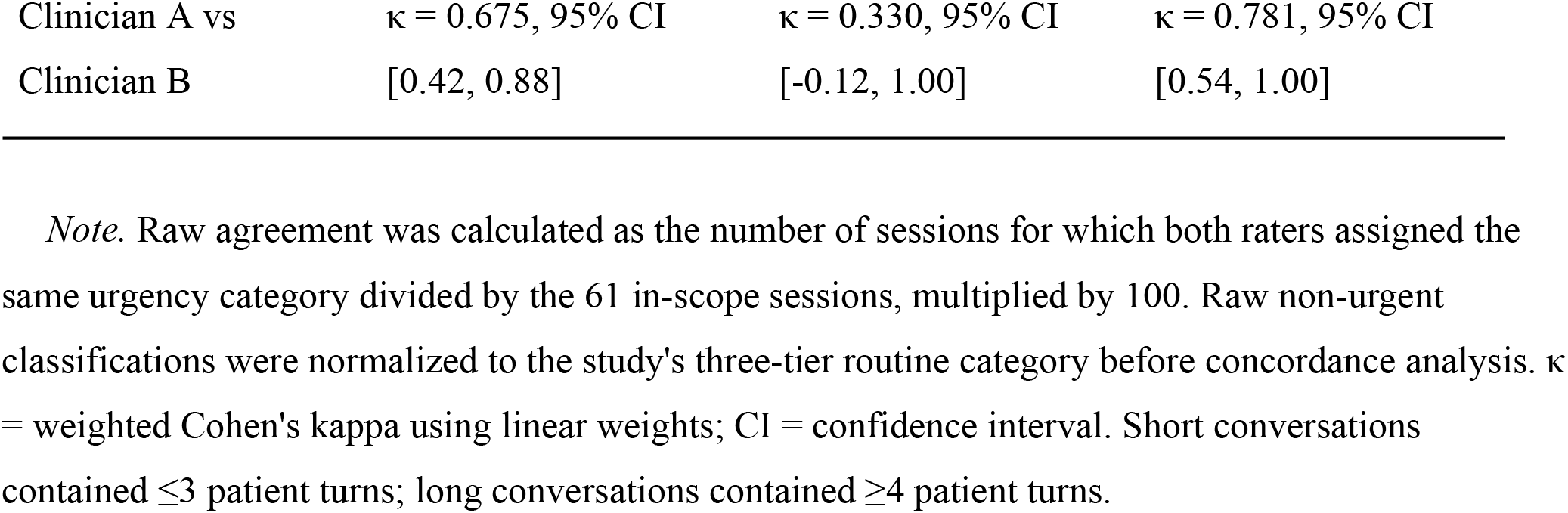
AI Triage Concordance by Conversation Length.

Agreement was stronger in longer conversations across all three pairings. Among conversations with four or more patient turns, weighted κ ranged from 0.705 to 0.781, compared with 0.330 to 0.646 among conversations with three or fewer turns. Confidence intervals were particularly wide in the short-conversation stratum (n = 23) and should therefore be interpreted cautiously.

Six of the 61 sessions showed disagreement between the two clinicians (S007, S014, S051, S057, S064, and S066). In five of these cases, Clinician A assigned the more cautious urgency classification; in one case, Clinician B assigned the more cautious classification. No third independent validator was available to adjudicate these disagreements.

### Subgroup comparison

No dimension showed a significant difference in access improvement between on-campus (n=15) and off-campus (n=27) participants after correction, a comparison underpowered at this sample size.

## Discussion

This study’s strongest finding is that MedGemma’s triage classifications, validated against two independent Nigerian clinicians, achieved raw agreement (90–93%) at or above the upper range reported for international symptom-checker tools (Wallace et al., 2022), and did not show the multi-turn degradation reported elsewhere (Fouda et al., 2026). To our knowledge, this is the first independent evaluation of MedGemma for AI-assisted triage among Nigerian university students. Notably, clinician-to-clinician agreement was itself substantial (κ=0.675), indicating the reference standard this study validated against was reasonably stable, a finding with direct implications for how future AI triage validation studies should be resourced: even where clinicians substantially agree, a second independent validator surfaces residual disagreement a single reviewer would hide entirely.

Access and equity outcomes were more modest. The consistent, if non-significant, directional improvement across most access dimensions is consistent with this study’s framing as feasibility-establishing rather than definitive (Murray et al., 2016); a sample of 42 paired participants is underpowered to detect anything but a large effect. Similarly, the subgroup comparison could not confirm or rule out a genuine access difference between on-campus and off-campus students at this scale.

Key limitations include: a single-institution, non-randomised design; a concordance sample size below what stable kappa estimation typically requires; no test of MedGemma’s performance in Nigerian languages or code-switched English; exclusion of participants with known disabilities, meaning the platform’s voice-interface accessibility rationale remains untested; and an achieved sample that departed from its planned recruitment balance because off-campus students self-selected at higher rates during the recruitment window. A larger, multi-institution, adequately powered follow-up, incorporating a third AI-validation reviewer and testing across Nigerian languages, is the clearest next step.

As registration occurred after data collection, this study cannot make the same guarantee against outcome-reporting bias that prospective registration provides; readers should weigh the pre-post access findings accordingly.

## Conclusion

This study demonstrates the technical feasibility, usability, and clinical concordance of a locally deployed, AI-assisted mobile health platform in a resource-constrained Nigerian setting, providing an independent validation of MedGemma for AI-assisted triage in a Nigerian university healthcare context. Access and equity outcomes were directionally positive but not statistically confirmed at this pilot scale, warranting a larger, adequately powered follow-up study.

## Data Availability

De-identified data supporting the findings of this study are provided as
Supplementary Data S1-S4, subject to applicable institutional data-protection requirements under Nigeria's Data Protection Act, 2023.

## Declarations

### Ethics approval and consent to participate

This study was approved by the University of Abuja Health Research Ethics Committee prior to data collection. Written informed consent was obtained from all participants.

### Consent for publication

Not applicable (no identifying individual data included).

### Clinical trial registration

This trial was registered with PACTR (PACTR202608826406831) on 28th July, 2026, after completion of data collection. Registration was not performed prospectively because the study was initially conducted as an institutional undergraduate research project without an a priori intention to seek external publication; retrospective registration was pursued once that intention was formed.

### Data availability

De-identified data supporting the findings of this study are provided as Supplementary Data S1–S4, subject to applicable institutional data-protection requirements.

### Author contributions

Ogorchukwu Emmanuel Ochem: Conceptualization, Methodology, Software, Formal Analysis, Writing – Original Draft Preparation. Okwuchukwu Praise Ezego: Data Curation, Formal Analysis (Statistical Validation), Writing – Review & Editing.

## Acknowledgements

The authors thank the student participants at the University of Abuja for their time and engagement throughout the pilot period. We also thank Dr Uzorchukwu Michael and Dr Idakwo Blessing for their independent clinical validation work under Objective 4, Prof. Ishaya M. Dagwa for supervision of the study and manuscript review, and the University of Abuja Teaching Hospital for institutional support during platform deployment and data collection.

## References

Alawode, G. O., & Adewole, D. A. (2021). Assessment of the design and implementation challenges of the National Health Insurance Scheme in Nigeria: a qualitative study among sub-national level actors, healthcare and insurance providers. BMC Public Health, 21(1), 124. 10.1186/s12889-020-10133-5

Cole, O. K., Abubakar, M. M., Isah, A., Sule, S. H., & Ukoha-Kalu, B. O. (2025). Barriers and facilitators of provision of telemedicine in Nigeria: A systematic review. PLOS Digital Health, 4(7). 10.1371/journal.pdig.0000934

Fouda, A. E., Hassan, A. A., Hanafy, R. J., & Fouda, M. E. (2026). PsychiatryBench: a multi-task benchmark for LLMs in psychiatry. Npj Digital Medicine, 9(1). 10.1038/s41746-026-02582-w

Google. (2026). MedGemma model card. In Health AI Developer Foundations.

Kruk, M. E., Gage, A., Joseph, N. T., Danaei, G., García-Saisó, S., & Salomon, J. A. (2018). Mortality due to low-quality health systems in the universal health coverage era: a systematic analysis of amenable deaths in 137 countries. The Lancet, 392(10160), 2203–2212. 10.1016/s0140-6736(18)31668-4

Murray, E., Hekler, E. B., Andersson, G., Collins, L. M., Doherty, A., Hollis, C., Rivera, D. E., West, R., & Wyatt, J. C. (2016). Evaluating digital health interventions: Key questions and approaches. American Journal of Preventive Medicine, 51(5), 843–851. 10.1016/j.amepre.2016.06.008

Ogueji, I. A., Ogunsola, O. O., Abdalla, N. M., & Helmy, M. (2023). Mistrust of the Nigerian health system and its practical implications: Qualitative insights from professionals and non-professionals in the Nigerian health system. Journal of Public Health, 32(2), 303–314. 10.1007/s10389-022-01814-z

Olatunji, T., Nimo, C., Owodunni, A., Abdullahi, T., Ayodele, E., Sanni, M., Aka, C., Omofoye, F., Yuehgoh, F., Faniran, T., Dossou, B. F. P., Yekini, M., Kemp, J., Heller, K., Omeke, J. C., Asuzu, C., Etori, N. A., Aimérou, N., Okoh, I., … Asiedu, M. (2024). AfriMed-QA: A Pan-African, Multi-Specialty, Medical Question-Answering Benchmark Dataset. In arXiv (Cornell University). Cornell University. 10.48550/arxiv.2411.15640

Penchansky, R., & Thomas, J. W. (1981). The Concept of Access. Medical Care, 19(2), 127–140. 10.1097/00005650-198102000-00001

Schmieding, M. L., Kopka, M., Schmidt, K., Schulz-Niethammer, S., Balzer, F., & Feufel, M. A. (2022). Triage Accuracy of Symptom Checker Apps: 5-Year Follow-up Evaluation. Journal of Medical Internet Research, 24(5). 10.2196/31810

Sim, J., & Wright, C. (2005). The Kappa Statistic in Reliability Studies: Use, Interpretation, and Sample Size Requirements. Physical Therapy, 85(3), 257–268. 10.1093/ptj/85.3.257

Stenberg, K., Hanssen, O., Edejer, T. T.-T., Bertram, M., Brindley, C., Meshreky, A., Rosen, J. E., Stover, J., Verboom, P., Sanders, R., & Soucat, A. (2017). Financing transformative health systems towards achievement of the health Sustainable Development Goals: a model for projected resource needs in 67 low-income and middle-income countries. The Lancet Global Health, 5(9). 10.1016/s2214-109x(17)30263-2

Wallace, W., Chan, C., Chidambaram, S., Hanna, L., Iqbal, F., Acharya, A., Normahani, P., Ashrafian, H., Markar, S. R., Sounderajah, V., & Darzi, A. (2022). The diagnostic and triage accuracy of digital and online symptom checker tools: a systematic review. Npj Digital Medicine, 5(1), 118. 10.1038/s41746-022-00667-w

